# Proof-of-concept pilot study on comprehensive spatiotemporal intra-patient heterogeneity for colorectal cancer with liver metastasis

**DOI:** 10.1101/2021.06.29.21259694

**Authors:** Ioannis D. Kyrochristos, Georgios K. Glantzounis, Anna Goussia, Alexia Eliades, Achilleas Achilleos, Kyriakos Tsangaras, Irene Hadjidemetriou, Marilena Elpidorou, Marios Ioannides, George Koumbaris, Michail Mitsis, Philippos C. Patsalis, Dimitrios H. Roukos

## Abstract

**Purpose:** The mechanisms underlying high drug resistance and relapse rates after multi-modal treatment in patients with colorectal cancer (CRC) and liver metastasis (LM) remain poorly understood. We evaluate the potential translational implications of intra-patient heterogeneity (IPH) comprising primary and matched metastatic intratumor heterogeneity (ITH) coupled with circulating tumor DNA (ctDNA) variability.

**Patients and methods:** According to our IPH-based protocol, 18 eligible patients with CRC-LM, who underwent complete tumor resection after neo-adjuvant treatment, with a total of 122 multi-regional tumor and perioperative liquid biopsies were analyzed via next-generation sequencing (NGS) of a custom 77-gene panel. The primary endpoints were the extent of IPH and the frequency of actionable mutations.

**Results:** The proportion of patients with ITH were 53% and 56% in primary CRC and LM respectively, while 35% of patients harbored de novo mutations in LM indicating spatiotemporal tumor evolution and the necessity of multiregional analysis. Among the 56% of patients with alterations in liquid biopsies, de novo mutations in cfDNA were identified in 25% of patients, which were undetectable in both CRC and LM. All 17 patients with driver alterations harbored actionable mutations, with an average of 3.2 oncogenic events per patient, for molecularly targeted drugs either approved or under evaluation in ongoing clinical trials or in pre-clinical studies.

**Conclusions:** Our proof-of-concept prospective study provides initial evidence and warrants the conduction of precision oncology trials to test the potential clinical utility of IPH-driven matched therapy.

## 1. INTRODUCTION

Dynamic evolution of genomic clones underlying cancer cell subpopulations and intratumor heterogeneity (ITH), as well as metastasis originating from tumor cells shed in the circulation and therapeutic resistance, represent the major causes of relapse and cancer-related death.^1-3^ The capacity of next generation sequencing (NGS) studies to identify multi-regional ITH and serial circulating cell-free DNA (cfDNA) or circulating tumor DNA (ctDNA) mutations responsible for intrinsic and acquired drug resistance has transformed cancer biology and translational research.^4,5^ We have developed and proposed a spatiotemporal concept of comprehensive intra-patient heterogeneity (IPH) with potential translation into Precision Oncology.^6^ In this pilot study we evaluate the translational efficacy of our IPH-based protocol to characterize and compare, for the first time, the ITH of primary colorectal cancer (CRC) and matched liver metastases (LM), in conjunction with the ctDNA mutational landscape. This holistic approach enables the detection of dynamic evolution of cancer genomes in time and space enabling the identification and potential targeting of all actionable mutations at different time points over the disease course.

Despite the widespread establishment of primary prevention, CRC remains the second leading cause of cancer-related death in industrialized western countries.^7^ In more than 50% of patients with CRC, the cancer metastasizes to the liver over the disease course, with half of the metastases being synchronous and half metachronous.^8^ To this day, liver resection remains the cornerstone of potentially curative treatment. However, only 15-20% of these patients are candidates for surgery aiming to complete tumor resection at diagnosis.^9^ Overall, treatment of colon cancer with resectable LM consists of surgery, upfront or after neoadjuvant chemotherapy, and adjuvant chemotherapy, while for rectal cancer, surgery after neoadjuvant chemoradiotherapy is the standard of care.^10^ A new addition to the guidelines is the option for neoadjuvant immunotherapy with nivolumab/ipilimumab or pembrolizumab in patients with high microsatellite instability (MSI), albeit based on limited data.^10^ For resectable metachronous metastases, upfront resection and adjuvant chemo is preferred over neoadjuvant treatment.^10^ Notably, no targeted agent has been approved for use in resectable disease. Nevertheless, recurrence-free survival for patients with resectable CRC-LM at 5 years remains only 30%, even after multimodal treatment.^9^

Over the past decade, an explosion in genome sequencing studies has provided accumulating data suggesting that a shift from single tumor biopsy to multi-regional tumor and liquid biopsy analysis could enable accurate genetic diagnosis to improve therapeutic decisions towards Precision Oncology.^11,12^ Established dynamic evolution of cancer genomes in time and space before and after treatment is reflected in cancer phenotypes, subclonal ITH and circulating plasma mutational variability.^5,13,14^ Indeed, this rapid progress is being translated into multiple underway clinical trials testing the efficacy of intratumor heterogeneity analysis and serial liquid biopsies to guide more effective individualized treatment.^15^ Considering the discovery of thousands of drug targets via NGS and genome editing technologies,^16,17^ IPH-matched therapy could maximize clinical benefit.^6,18^ Based on intra-lesion and serial ctDNA variability, comprehensive patient-specific tumor and ctDNA analysis could empower the optimization of decision-making on the selection of targeted drugs.^13^

To assess ITH and serial ctDNA mutational heterogeneity in the perioperative setting, we designed a prospective protocol encompassing multiple intra-lesional and matched plasma samples for each individual patient. We enrolled patients with CRC and LM who underwent resection of the primary and metastatic tumors with curative intent, after neo-adjuvant treatment. The comparisons between primary, metastatic and plasma mutational variability can dissect the dynamic evolution of genomic clones in time and space, orchestrating individual cancer phenotypes and drug resistance. Therefore, the concept of IPH proposed in our pilot study could enable the shift from single tumor to multiple tumor and liquid biopsy sampling, potentially improving diagnostic guidelines, and providing translational implications for personalized novel drug combinations.

## 2. PATIENTS AND METHODS

A total of 28 patients diagnosed with metastatic colorectal adenocarcinoma were treated in our surgical department in the University Hospital of Ioannina between January 2017 and December 2019 and were enrolled in this study. All patients signed a consent form for analysis of biomaterials provided by our institution’s ethics committee. After initial histopathological quality control for adequacy of both primary and metastatic tumor tissue 10 patients were excluded due to inadequate tissue availability from the primary, metastatic or both sites. Thus, 18 patients were included in our final analysis. For anonymization purposes, a unique code was assigned to each patient (AA to AR). In our cohort, 13 patients had synchronous and the remaining 5 metachronous liver metastases (LM). All patients were previously subjected to neo-adjuvant chemotherapy, according to the recommendations of our institution’s Multidisciplinary Tumor Board. For patients who were subjected to primary tumor resection before the initiation of the study, multi-regional primary tumor (PT) samples were retrospectively collected in our Department of Pathology. In total, 94 FFPE samples were collected from both PTs and LMs. Eighty-six out of 94 FFPE samples passed quality control (QC) requirements (DNA yield, DNA quality) to be further subjected to downstream analysis. For 16 out of 18 patients, plasma was collected at multiple time points during treatment, before and after surgery, to assess the molecular dynamics of the disease using liquid biopsies. A total of 38 plasma samples were collected out of which 36 samples passed QC requirements. Overall, out of 132 samples in total, data were collected and analyzed for 122 samples (92.4% QC success). A flowchart summarizing our study population and sample analysis is delineated in Figure 1.

**Figure 1:**
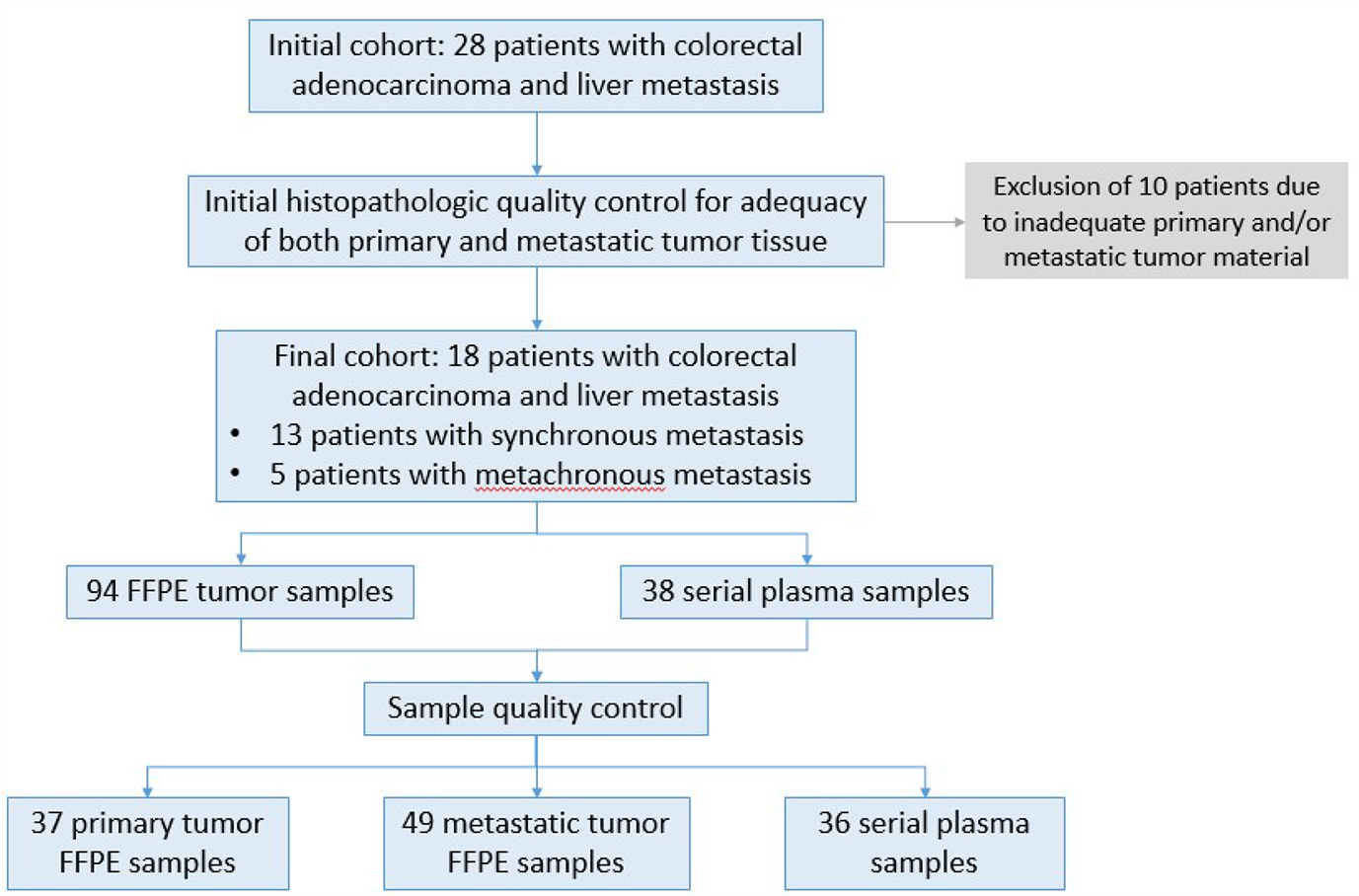
Description of patient samples. Twenty-eight patients diagnosed with colorectal cancer and liver metastasis were enrolled in the study. After initial histopathologic quality control for adequacy of tumor tissue 18 patients were subjected to further analysis. For 16 patients both FFPE tissue as well as blood samples were collected at different time points from diagnosis, before and after surgery and during treatment and monitoring. For two patients FFPE samples were available from different sites of the primary and metastatic site. A total of 122 samples passed QC requirements (92.4%) for NGS analysis.

### DNA preparation

DNA was extracted from FFPE tissue sections using the GeneRead DNA FFPE Kit (Qiagen) following the manufacturer’s instructions. DNA was quantified with a fluorometric based assay for FFPE tissue-derived DNA (Qubit flex fluorometer, Qubit dsDNA high sensitivity assay, Thermo Scientific). ctDNA was extracted from 4ml plasma using the QIAamp Circulating Nucleic Acid Kit (Qiagen) following the manufacturer’s instructions.

### Library preparation, enrichment, and sequencing

DNA libraries from FFPE and plasma samples were prepared using established protocols. Evaluation of library samples was performed using the 2200 Agilent Tapestation system (D1000 ScreenTape, Agilent). DNA enrichment for the genomic regions of interest was carried out using an in solution-hybridization based method using TACS (TArget Capture Sequences) specifically designed to capture selected loci in the genes of interest. TACS were then immobilized on streptavidin-coated magnetic beads for subsequent hybridization with the DNA libraries. A custom NIPD Genetics tumor profile gene assay was used for the identification of single nucleotide variants (SNVs), small insertions and deletions (indels), copy number alterations (CNAs) and rearrangements and microsatellite instability (MSI) detection (Table 1). Eluted samples were amplified using outer-bound adaptor primers. Enriched DNA libraries were then normalized and subjected to sequencing on an Illumina sequencing platform.

**Table 1:** Custom 77-gene panel for targeted next-generation sequencing analysis

| Single Nucleotide Variants (SNVs) / Insertions and Deletions (Indels) (67 genes) | Copy-Number Alterations (17 genes) | Translocations (10 genes) |
| --- | --- | --- |
| AKT1, ALK, APC, AR, ARAF, ATM, ATRX, BARD1, BRAF, BRCA1, BRCA2, BRIP1, CDH1, CDKN2A, CHEK2, CIC, CTNNB1, DDR2, DICER1, EGFR, ERBB2, ERBB3, ERBB4, ESR1, FBXW7, FOXA1, FOXL2, FUBP1, GATA3, GNA11, GNAQ, GNAS, H3F3A, IDH1, IDH2, JAK2, KEAP1, KIT, KRAS, MAP2K1, MAP3K1, MET, MLH1, MRE11A, MSH2, MSH6, MTOR, NBN, NF1, NRAS, NTRK1, PALB2, PIK3CA, PIK3CB, PMS2, POLE, PTEN, RAD51C, RAD51D, RAF1, RET, RUNX1, SMAD4, SPOP, STK11, TERT, TP53 | AR, CDKN2A, EGFR, ERBB2, ESR1, FGFR1, FGFR2, FGFR3, KIT, KRAS, MET, MYC, MYCN, PIK3CA, PTEN, RB1, TP53 | ALK, BRAF, FGFR3, NF1, NTRK1, NTRK2, NTRK3, RET, ROS1, TMPRSS2 |

\*Includes MSI assessment

### Bioinformatics analysis

#### Tissue biopsy analysis pipeline

Sequencing data were de-multiplexed with bcl2fastq (v.2.16.0) and paired-end DNA sequencing reads were processed to remove adapter sequences and poor-quality reads. The remaining sequences were aligned to the human reference genome build (hg19) using the Burrows-Wheeler alignment algorithm.^19^ Duplicate read entries were removed^20^ and aligned reads files were converted to a binary (BAM) format. Variant calling was performed using a versatile somatic variant caller.^21^ All detected variants were filtered, annotated and classified based on well known, publicly available, disease databases (COSMIC18 and ClinVar.19). Benign or likely benign variants were filtered out. Only variants with strong or potential clinical significance according to AMP/ASCO/CAP guidelines (TierI/TierII) were reported for each tested sample (20). Gene-level Copy Number Variants (CNVs) were detected using an in-house bioinformatics pipeline that implements a circular binary segmentation method (ref4).^22^ Translocation calling is performed by utilizing discordant pair and split-read alignments following local assembly, realignment and an in-house filtering pipeline to refine the set of candidate events.^23-26^

#### cfDNA analysis pipeline

Sequencing data were de-multiplexed with bcl2fastq (v.2.16.0) and paired-end DNA sequencing reads were processed to remove adapter sequences and poor-quality reads. The remaining sequences were aligned to the human reference genome build (hg19) using the Burrows-Wheeler alignment algorithm. Duplicate reads were identified, grouped by their Unique Molecular Identifier (UMI) family and processed to produce consensus reads per UMI family (fgbio). Allelic count information for all targeted loci was used to calculate the variant allele frequency for each substitution and short insertion/deletion. A statistical error-correction model (at a base-pair resolution) was subsequently applied to refine the set of positive variant calls. Variant annotation and classification was performed using the Varsome Clinical platform. Translocation calling was performed by utilizing discordant pair and split-read alignments following local assembly, realignment and a filtering pipeline to refine the set of candidate events. Transformed read depth information on pre-defined genomic windows spanning the regions of interest were normalized utilizing a, multistep statistical method (applying a within- and between-samples normalization approach). Normalized read depth data were processed to detect copy number changes.

## 3. RESULTS

Based on our analysis, mutations were detected in 17/18 (94.4%) of patients in our cohort, and the number of variants per patient ranged from 0 to 18. Overall, mutations in 28 genes were identified, adding up to an average 4.7 variants per patient. In agreement with published literature, the most frequently mutated genes were *APC* and *TP53*, followed by *PIK3CA* and *KRAS*, while 3 or fewer variants were identified for all other genes. Importantly, the majority of identified variants (19/28, 69%) were rare and detected in only one patient. Moreover, a significant proportion of variants was identified in potentially clinically relevant genes not routinely tested in day-to-day practice. The majority (60%) of alterations regarded missense mutations, while other alteration types included frameshift (14.1%) and nonsense (11.8%) mutations, amplifications (8.2%) and others. Figure 2 summarizes our findings on the frequency of mutated genes, mutation types and patient variants.

**Figure 2:**
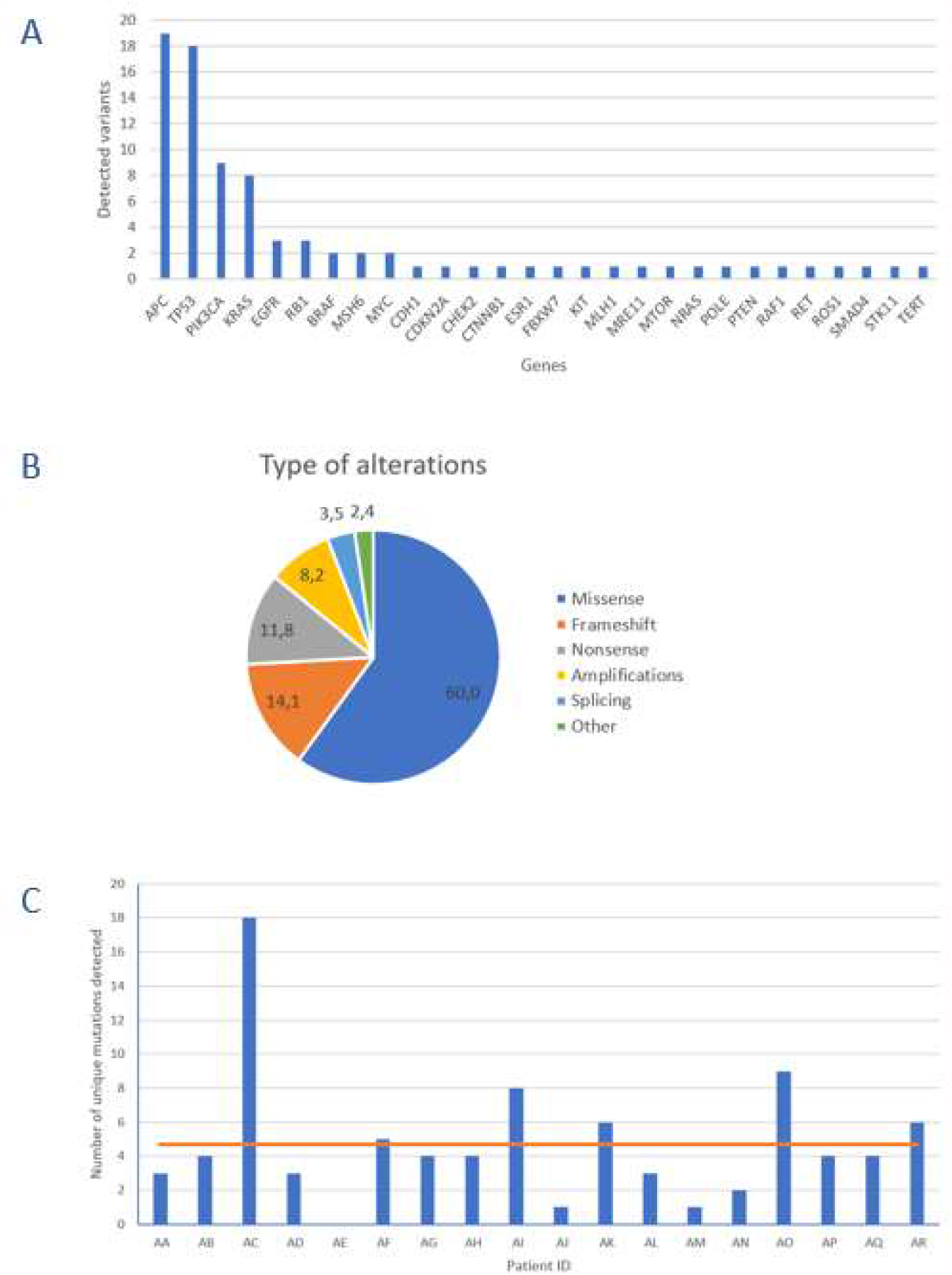
Overview of alterations observed in our study. A. Frequency of mutations observed per gene: The frequency of mutations is estimated based on the cumulative list of variants per patient. The number of variants identified in all patients is shown. B. Distribution of different types of genetic alterations identified in the study. C. Total number of variants per patient in the cohort.

### Intra-tumor heterogeneity

Initially, we aimed to detect and characterize the extent of ITH within matched primary and metastatic lesions. Concerning primary tumor analysis, we analyzed 2-4 multiregional PT samples from 15 patients, while regarding liver metastases, we analyzed 2-5 multiregional LM samples in each of 16 patients. For one patient (AG), we analyzed two distinct liver metastases, one synchronous and one metachronous. Due to quality control exclusion, three patients had single PT and/or LM samples. There was no PT sample available for one patient. In our cohort, increased number of multiregional samples did not correlate with the frequency of observed ITH as compared to dual samples in primary or metastatic tumors, although our relatively small cohort sample warrants further large-scale investigation. Quite notably, 53% (8/15) of patients with multi-regional primary biopsies featured ITH between spatially distinct geographical regions of the PT at variable degrees, with homogeneity of observed variants between regions ranging from 83% to under 15%. Similarly, variable multi-regional ITH of the LMs was observed in 56% (9/16) of patients between different regions of the same metastasis. These results indicate that analysis of multiple spatially distinct samples is meaningful in more than half of lmCRC patients, with potentially significant clinical implications as discussed below.

### Circulating tumor DNA variability

Based on the highly compelling potential for a non-invasive blood-based patient monitoring strategy, we additionally analyzed 36 serially collected plasma samples before and after therapy from 16 of 18 patients utilizing NIPD Genetics custom gene assay, to explore the temporal dynamics of ctDNA variability. Plasma ctDNA analysis identified mutations in 60% (9/15) of patients with both pre- and post-operative plasma samples. Tumor mutations were detected in the ctDNA analysis in 8 and 2 of 16 patients pre- and post-operatively respectively. There was a clear tendency for pre-operative tumor mutations in plasma to be undetectable in ctDNA after complete tumor resection (R0 surgery) with curative intention, indicating the well-established association of plasma mutation detection and tumor burden. Persistence of ctDNA mutations post-operatively, as for example in patient AA for whom a staged hepatectomy was planned but not performed, thus, subjected to R0 resection, correlated with early relapse (<1 year) and adverse oncological outcome.

### Comprehensive intra-patient heterogeneity

The first most ambitious aim of our project was to dissect the comprehensive spatiotemporal genetic IPH for each individual patient, comprising the ITH of primary and metastatic lesions, as well as temporal ctDNA mutational heterogeneity. Most importantly, genetic differences between matched PT and LMs, meaning the presence of mutations in either the primary or metastatic tissue but not in both, were detected in 53% (9/17) of cases with both PT and matched metastatic tumor (MT) samples, with 35% (6/17) of patients harboring de novo mutations present only in LMs, indicating dynamic clonal evolution. Overall, 41% of variants observed in solid tumor samples were ubiquitously shared by all regions between primary and liver lesions. These results suggest that multiple biopsies of both primary and matched metastatic lesions are required to delineate cancer genetic diversity with potentially crucial implications for the clinic. Quite impressively, ctDNA analysis effectively dissected the complete variability of PTs and matched MTs in 25% (4/16) of cases, identifying all intra-tumorally identified mutations in these patients, suggesting the potential use of liquid biopsy for delineating ITH.. Lastly, in 4 of 16 patients (patients AA, AD, AG, AJ), ctDNA analysis uncovered mutations, which were not detected in the primary or metastatic tumors and could represent the result of dynamic subclonal evolution, which needs further investigation. Notably, putatively actionable mutations identified exclusively in liquid biopsies, including KRAS, *CHEK2, PIK3CA, TP53* and others, could provide important translational therapeutic implications. All of the above support our hypothesis that a holistic approach to the oncological patient requires rigorous combinatorial analysis of spatiotemporally collected primary tumoral, metastatic and plasma samples to improve the accuracy in characterizing the complete tumor genetic diversity.

### Intra-patient mutational heterogeneity-driven targeted therapy

The second major aim of our study was to identify potential oncotargets and characterize their clonality within PT and matched LMs, in order to clarify whether a single biopsy could effectively guide therapeutic decision-making. In our cohort, all patients presented one or multiple therapeutic opportunities specifically for targeted treatment. Of the 18 patients, 9 (50%) were wild-type for *KRAS*/*NRAS* and could derive potential benefit from EGFR inhibitors, such as cetuximab and panitumumab. Two patients had *BRAF* mutations, presenting an opportunity for treatment with *BRAF*-inhibitors, already approved for use in non-resectable CRC. Moreover, 50% (9/18) of participants had variant targets of drugs already approved for other cancer types, suggesting a potential benefit through drug repurposing. Additionally, 72% (13/18) and 67% (12/18) of patients could be matched to targeted drugs under clinical or pre-clinical evaluation respectively, in CRC or other cancer types. It is worth noting that, out of 19 detected rare variants, 10 were potentially actionable. Collectively, 94% (17/18) of participants in our cohort featured targets of not-yet-approved agents in the clinical or pre-clinical stages of development.

A notable example is patient AC, a patient with established Lynch syndrome according to the Amsterdam criteria, as well as microsatellite instability previously identified using a panel of five markers (BAT-25, BAT-26, D5S346, D17S250 & D2S123) through PCR-based testing and confirmed by our assay. Our analysis also confirmed a mutated *MSH6* gene in all intratumor samples as well as its presence in plasma. In this patient, more than 10 putatively actionable variants were identified, with the majority being subclonal, suggesting the potential for extensive drug combinations, including immunotherapy, guided by intratumor and circulating DNA heterogeneity.

A crucial parameter hindering the potential therapeutic utilization of genetic findings is the putative spatial and temporal subclonality of actionable mutations, which, if validated, could not be addressed via a single tumor biopsy. Indeed, our study identified an average of 3.2 potentially actionable variants per patient, suggesting the capacity for combinatorial targeted therapy, following large-scale validation. However, based on comparative primary and metastatic tumor analysis, 71% (12/17) of patients with detectable mutations harbored subclonal putative oncotargets not identified in all tumor samples, which could potentially be masked by single-biopsy analysis (Figure 3).^27,28^ Analysis of ctDNA uncovered potentially druggable variants in all nine patients with detectable plasma mutations, indicating the potential role of ctDNA in guiding therapeutic decisions following validation. Quite notably, 33% and 22% of participants harbored putatively actionable variants only in LM and plasma samples respectively, which would not have been identified by PT analysis alone, although our study is limited in discovering plasma-exclusive variants, as described below. Overall, almost 65% of all oncotargets detected in our cohort featured diverse degrees of intratumor and circulating variability among samples from the same patient. Therefore, our study strongly supports the hypothesis that therapeutic decision-making should not be limited to single PT samples and warrants further evaluation of spatiotemporally collected samples from matched PTs, MTs and plasma in large-scale studies to establish the clinical utility of comprehensive IPH. A summary of our findings on genetic spatiotemporal heterogeneity in tumor and liquid samples is delineated in Figure 4. In addition, most crucial findings are explained in Box 1.

**Figure 3:**
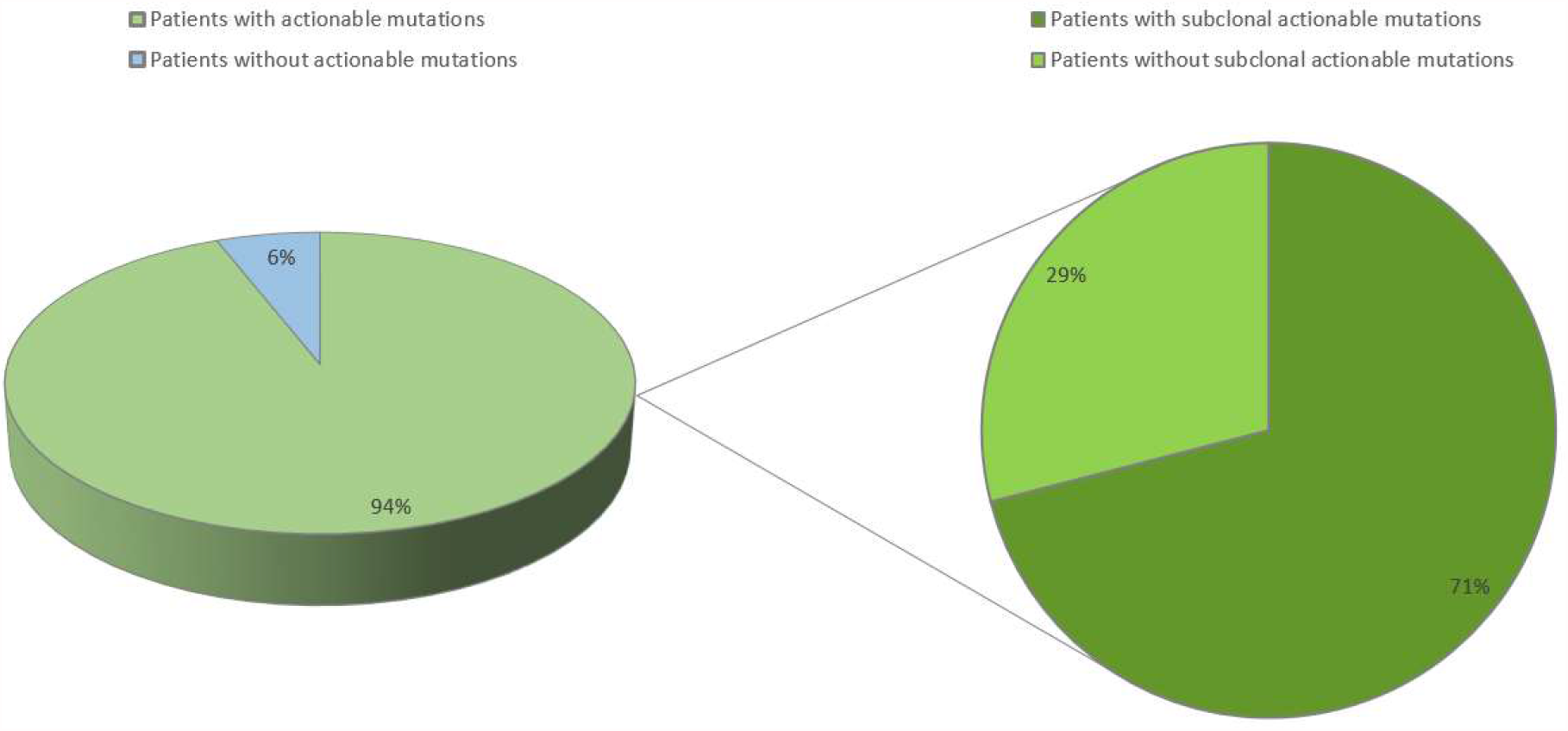
Overall detection of potentially actionable mutations and their clonal status in our study.

**Figure 4:**
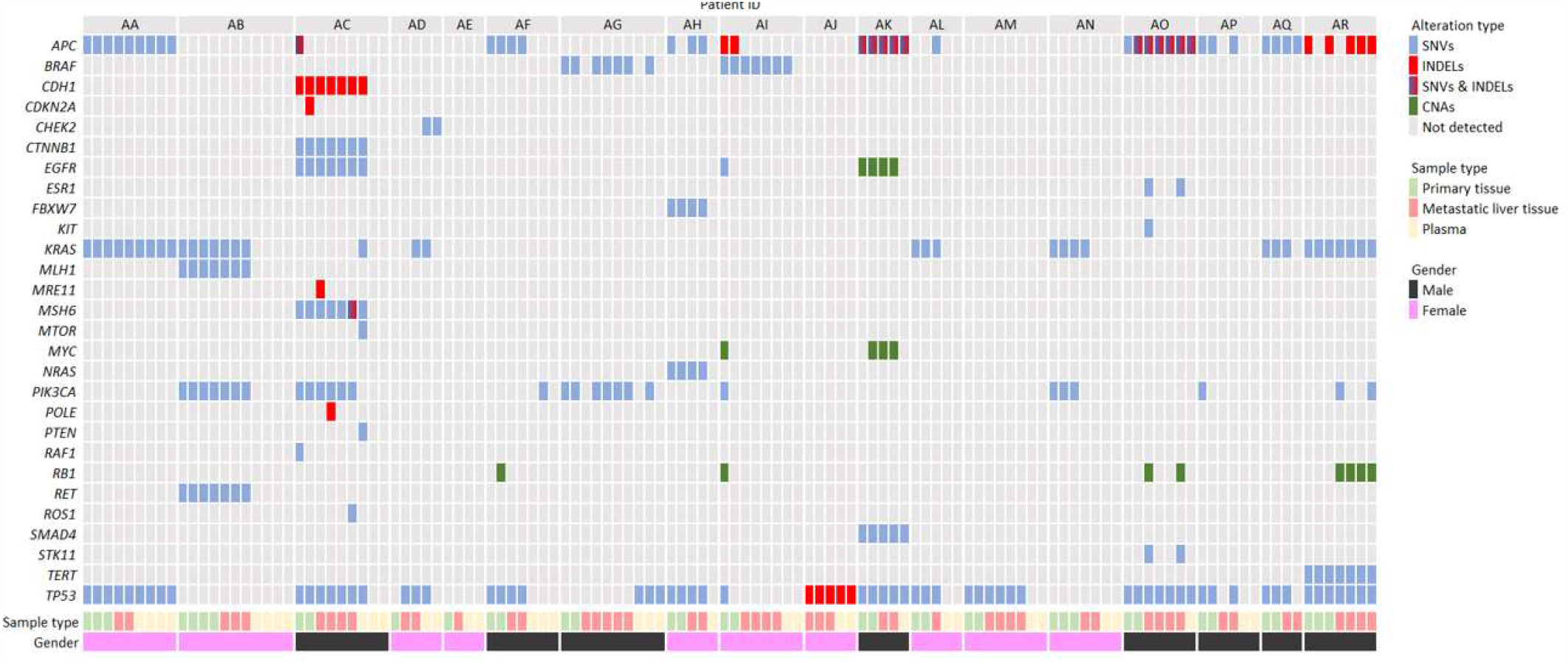
Summary of the distribution of genetic alterations among all tissue and liquid biopsy samples from our cohort of 18 patients with metastatic colorectal adenocarcinoma.

## 4. DISCUSSION

Our proof-of-concept pilot prospective study on comprehensive intra-patient genetic heterogeneity in patients with resected CRC and LM provides new diagnostic, predictive and therapeutic implications. In the present cohort, 53% and 56% of patients harbored ITH of the primary and metastatic tumors respectively, while 53% had genetic differences between matched primary and metastatic samples. Combining both primary and metastatic ITH with matched plasma cfDNA, 25% of patients had aberrations in plasma but not in tumor specimens which highlights the potential benefit of ctDNA analysis in capturing tumor heterogeneity which might be missed from analysis of only a few FFPE sections that offer solely a snapshot of the tumor’s molecular profile. Almost all patients had cancer targets for approved drugs and/or agents under investigation in ongoing clinical trials^15^ or pre-clinical studies. Our data are consistent with multiple genomic and transcriptomic studies on spatial and temporal dynamic evolution of cancer genomes,^27,29^ underlying intratumor subclonal cell populations.

Multiple genomic studies have identified ITH as an integral part of cancer evolution in solid tumors.^28,30^ This multi-regional variability has been strongly correlated with intrinsic and acquired drug resistance and relapse.^27,31,32^ Indeed, consistent with our work, several studies have uncovered extensive ITH of primary CRC as a prognostic factor for metastasis, as well as a predictor of drug resistance.^33-35^ With regards to ITH of the liver metastasis, our study is among the very few published reports with a strict protocol, enabling the exact identification of metastatic genetic ITH in 56% of patients. Most available studies have evaluated the variability between matched primary and metastatic lesions through either single or multi-regional samples. Based on a single biopsy, data remain controversial regarding the degree of heterogeneity between primary and matched metastatic lesions.^36-38^ Similarly, intra-lesion sampling of both primary CRC and LM has provided contradictory findings. For instance, Siraj et al. demonstrated high genomic concordance between primary CRC and matched LM via whole-exome sequencing of a total of 191 samples.^39^ By contrast, Hu and colleagues, in a whole-exome sequencing cohort consisting of 118 biopsies from 23 patients, uncovered extensive inter- and intra-lesion heterogeneity,^40^ in accordance with our prospective cohort. Whether a shift from the current standard of single-tumor biopsies to multi-sampling from matched primary and metastatic lesions can enable more accurate diagnosis and decision-making on novel combinations of molecularly targeted drugs remains at the present unclear. By identifying putatively targetable mutations in nearly all patients, of which approximately 65% featured variable intratumor and ctDNA heterogeneity, our pilot study strongly supports the prospective evaluation of this concept within Precision Oncology trials. Indeed, the proportion of patients with clinically actionable mutations in recent large-scale consortia based on single-biopsy NGS, such as The Cancer Genome Atlas and the Pan-Cancer Analysis of Whole Genomes, ranged between 57% and over 75%,^17,41^ highlighting that multi-sampling could substantially increase the discovery rates of cancer targets.

Targeted NGS, whole-exome sequencing and whole-genome sequencing of cfDNA or ctDNA, as well as analysis of circulating tumor cells, are receiving tremendous attention towards implementation into the clinical setting for early diagnosis, individualized prediction of drug response, patient monitoring to readily detect relapse and drug development.^29,42^ Indeed, beyond large innovative projects,^43^ several comprehensive pan-cancer multi-gene panels, including for CRC, have already been developed and approved by federal regulatory institutions as companion diagnostics for tumor profiling within modern guidelines. Those most notably include the FoundationOne Liquid CDx assay that includes >300 genes as well as measurements of tumor mutational burden (TMB), MSI and tumor fraction values,^44,45^ the MSK-IMPACT and MSK-ACCESS panels for tumor and plasma respectively, analyzing 468 and 129 cancer genes,^45-47^ the Guardant360 CDx, which includes 54-73 genes and detection of MSI,^45,48^ as well as multiple other platforms commercially available or under development.^29^ On this basis, the promising findings provided via our custom 77-gene panel could be incorporated into the clinical setting, following large-scale validation. Additionally, completed early-phase clinical trials, such as the TARGET^49^ and the I-PREDICT^50^ studies have demonstrated potential clinical benefit from liquid biopsy-guided drug target detection and molecularly matched therapy.

Based on our recent published work on the comprehensive model of IPH,^6^ in the present study, we have explored the potential translational implications of IPH, comprised by the ITH of primary and metastatic lesions in combination with plasma DNA mutational landscapes. This integrated framework has highlighted the necessity for multiple sample analysis from both tumor and ctDNA in the perioperative setting. This approach could potentially transform decision-making on neoadjuvant or adjuvant treatment following complete tumor resection to overcome the unmet clinical challenge of substantial therapeutic resistance and relapse rates among patients with resectable colorectal cancer with liver metastasis. In fact, 35% of patients in our cohort harbored de novo mutations in the metastases, potentially unraveling the capacity to improve therapeutic decisions. Moreover, de novo mutations were quite impressively identified in the ctDNA infrom 25% of patients but not in the multi-regional analysis of primary and metastatic tumors. However, this finding requires further evaluation via DNA analysis of white blood cells to differentiate cancer from germline mutations or potential clonal hematopoiesis.^51^ Additionally, on the basis of WGS on over 2,500 cancer samples, the PCAWG initiative identified actionable mutations in 60% of tumors,^30^ while in our integrated analysis of both multi-regional tumor and serial liquid biopsies, all patients with detectable mutations harbored actionable events, supporting the translational framework of detailed tumor and plasma analysis. Nevertheless, as demonstrated by our results, isolated analysis of primary CRC, metastatic tissue or ctDNA is unable to uncover the complete mutational landscape for each individual patient, highlighting the translational importance or our work.

Despite intriguing findings, our study is presented with several limitations. First, the number of enrolled patients and total samples analyzed is relatively small to extract definitive conclusions. Second, our plasma analysis is lacking in serial postoperative samples over the course of disease to evaluate the potential for early relapse detection and potential therapeutic targeting. And third, our analysis focuses on the detection of actionable mutations and matched targeted drugs, while not exploring putative immunotherapeutic implications. It should however be noted that our analysis was a pilot study, which was designed to explore the feasibility and potential clinical applications of the IPH concept, therefore, encouraging further extensive work and large-scale prospective studies and precision clinical trials.

### Future perspectives

Currently, ongoing projects and underway clinical trials are evaluating the clinical utility of cancer type-specific ITH and NGS of plasma DNA mutational heterogeneity, as well as tumor-infiltrating immune and stromal cells of the tumor microenvironment, to establish Precision Oncology and Immunology in the clinical setting.^12,15,52,53^ Expanding our holistic IPH approach to include intratumor interactions between cancer and environmental immune cells, pioneering single-cell genome sequencing, editing and machine learning technologies, as well as liquid biopsies of peripheral blood for cfDNA, circulating tumor and immune cell analysis raise novel expectations to realize patient-specific optimal precision immuno-oncological treatment.^18,29,54-58^

## 5. CONCLUSIONS

Our IPH-based concept with a pilot-level tNGS analysis of 122 multi-regional tumor and liquid biopsies with a custom 77-gene assay has provided initial evidence on the necessity of multiple spatiotemporal sampling in patients with resectable CRC with LM. This region-to-region NGS analysis of primary and matched metastatic lesions, as well as perioperative plasma samples, has enabled the dissection of primary and metastatic ITH and ctDNA variability. Indeed, there was substantial heterogeneity between primary, metastatic and circulating mutational landscapes, unraveling the dynamic evolution of cancer genomes, underlying tumor progression, metastasis, drug resistance and relapse. Our data uncover the significance of ITH and cfDNA as predictors of response to molecularly targeted drugs. Moreover, the identification of clinically actionable mutations by ctDNA analysis highlights the importance of evaluating tumor dynamics and heterogeneity using liquid biopsy to guide therapy selection and better stratification of patients for clinical trial enrollment and treatment-response evaluation. The holistic approach in our work detected in average 3.2 actionable events per patient, warranting the conduction of precision clinical trials to assess the clinical utility of novel targeted drug combinations in the adjuvant or neo-adjuvant setting. In summary, comprehensive IPH-based translational research and clinical trials are expected to transform current treatment guidelines towards the implementation of Cancer Precision Medicine in the clinical setting, to overcome the unmet challenges of high drug resistance and relapse rates.

## Data Availability

All relevant data are included in this submission.

---

**Box 1:** Summary of the most crucial findings in our prospective cohort

|  |  |  |
| --- | --- | --- |
| ITH of the primary tumor | The proportion of patients harboring variable levels of ITH of the primary colorectal tumor | 8/15 patients with multi-regional samples from the primary tumor, 53% |
| ITH of the liver metastasis | The proportion of patients harboring variable levels of ITH of the liver metastatic lesion | 9/16 patients with multi-regional samples from liver metastases, 56% |
| Genetic heterogeneity between primary and matched metastatic tumors | The proportion of patients harboring mutations detected either in the primary or the matched metastatic tumor but not in both | 9/17 patients with matched primary and metastatic tumor samples, 53% |
| De novo mutations in liver metastases | The proportion of patients harboring de novo mutations in liver metastases not found in the primary tumor, including potentially actionable variants indicating dynamic clonal evolution | 6/17 patients with matched primary and metastatic tumor samples, 35% |
| Detection of cfDNA mutations | Proportion of patients in which tumor mutations were detected in cfDNA pre-operatively | 8/16 patients, 50% |
|  | Proportion of patients in which tumor mutations were detected in cfDNA post-operatively | 2/16 patients, 12.5% |
| Potentially actionable mutations | 9/18 (50%) and 17/18 (94%) patients could benefit from repurposing of already approved drugs and agents under clinical or pre-clinical development respectively. Overall, an average of 3.2 actionable mutations per patient were identified. | 12/17 (71%) patients with detectable mutations harbored potentially actionable alterations not ubiquitously shared by all tumor samples, indicating the need for spatiotemporal sampling to increase therapeutic accuracy |

